# Retrospective Analysis of Interventions to Epidemics using Dynamic Simulation of Population Behavior

**DOI:** 10.1101/2020.11.09.20228684

**Authors:** Jenna Osborn, Shayna Berman, Sara Bender-Bier, Gavin D’Souza, Matthew Myers

## Abstract

Retrospective analyses of interventions to epidemics, in which the effectiveness of strategies implemented are compared to hypothetical alternatives, are valuable for performing the cost-benefit calculations necessary to optimize infection countermeasures. SIR (susceptible-infected-removed) models are useful in this regard but are limited by the challenge of deciding how and when to update the numerous parameters as the epidemic changes in response to population behaviors. Behaviors of particular interest include facemasks adoption (at various levels) and social distancing. We present a method that uses a “dynamic spread function” to systematically capture the continuous variation in the population behavior, and the gradual change in infection evolution, resulting from interventions. No parameter updates are made by the user. We use the tool to quantify the reduction in infection rate realizable from the population of New York City adopting different facemask strategies during COVID-19. Assuming a baseline facemask of 67% filtration efficiency, calculations show that increasing the efficiency to 80% could have reduced the roughly 5000 new infections per day occurring at the peak of the epidemic to around 4000. Population behavior that may not be varied as part of the retrospective analysis, such as social distancing in a facemask analysis, are automatically captured as part of the calibration of the dynamic spread function.

## 1. Introduction

Retrospective analyses of strategies used to contain epidemics such as COVID-19 are valuable for countering successive waves of the infection, selecting countermeasures for future epidemics, and educating the population regarding the efficacy of implementing behavioral modifications. In particular, public-health agencies responsible for recommending types of personal protective equipment (PPE) to stockpile in anticipation of a future epidemic can benefit from the cost-benefit information yielded by retrospective analyses. Mathematical models, including those of the SIR type, can be helpful in providing a quantitative framework for the analyses. SIR models have been applied during the COVID-19 pandemic (Stutt et al. 2020, Giordano et al. 2020, Cooper et al. 2020, Bertozzi et al. 2020), primarily in a predictive capacity. Some of the studies (Ngonghala et al. 2020, Eikenberry et al. 2020) have predicted the infection dynamics for different intervention strategies, using a specific infection scenario (e.g. New York State).

A formidable challenge in applying SIR models is prescribing the values of the numerous parameters, and updating them to simulate evolving infection dynamics, as population behaviors (such as facemask adoption and social distancing) change in response to interventions. Typically, behaviors will change in a continuous manner rather than abruptly, for example, the gradual adoption of face masks by an affected population. Such gradual changes are difficult to capture in SIR models by periodically adjusting the parameters manually. The challenge is further accentuated by the high sensitivity of the predictions to some of the parameter values (Giordano et al. 2020). Often, parameter choices are based upon best guesses, or closeness of fit (sometimes visual) of computed profiles with published curves (Cooper et al. 2020).

In this paper, we introduce a modification of traditional SIR models that incorporates a “dynamic spread” function that captures changes in population behavior in a continuous manner. There is no need to adjust parameters manually as interventions are implemented during the course of the infection. The dynamic spread function satisfies a differential equation with variable coefficients. These coefficient functions are obtained from a calibration procedure employing the published infection profile for the region of interest. The computed dynamic spread function reproduces the infection profile resulting from the baseline intervention strategy implemented over the course of the epidemic. Subsequently, the spread function can be systematically modified to analyze the effect of alternate intervention strategies. We illustrate the process using the COVID-19 crisis in New York City (CNYC) and New York State. The reduction in infection rate realizable in CNYC from alternative intervention strategies, including increased levels of mask usage and deployment of masks with higher levels of filtration, is estimated.

## 2. Methods

We illustrate the technique using a 4-equation SIR model (Stilianakis and Drossinos 2010 Myers et al. 2016). The evolution of the susceptible, infected, and removed populations is simulated, as is the droplet transmission. The model assumes that the infection dynamics are dominated by one transmission mode (e.g. airborne particulates), and the parameter values are appropriate for all particle sizes contributing to that mode (though interpretation of the resulting equations as an average for a broad particle distribution is possible (Myers et al. 2016). More complicated SIR models can be useful, particularly if it is desired to model the details of the infection dynamics, e.g. quantifying the roles played by symptomatic and asymptomatic individuals (Stutt et al. 2020). Our intention is to use the simplest model that can capture the baseline population behaviors and vary the critical ones retrospectively, with the hope that the model can be understood and used by non-experts such as policy makers. Additionally, as noted by Siegenfeld et al. (2020), simpler models can prove more useful than complex ones, in part because accurate data is often not available to inform complicated formulations. Finally, we expect that some of the technique we present can be extended to more complex models.

### 2.1 Overview of Strategy

The dynamic-spread function is the critical element of a systematic procedure for re-purposing SIR models to perform retrospective studies. The 5 steps in the procedure are listed below and implemented subsequently.

1. Use the rate of change (measured by the number of new infections per day), *dS/dt*, of the susceptible population *S*, as the primary dependent variable. The derivative profile, which we call *T(t)*, does not require the number of recovered patients to be tracked.
2. Normalize variables and identify critical dimensionless parameters. Formulating the model in terms of dimensionless clusters of parameters reduces the number of independent quantities that must be prescribed to run the model, and aids in identifying the most critical parameters.
3. Allow the dimensionless parameter *δ*, which contains the product of the infection transmission rate and the virus production rate, to vary with time, and account for its time dependence in the governing differential equation for *T(t)*. We denote *δ(t)* the “dynamic-spread” function, as it contains the elements that both vary with time and govern the rate of spread of the infection. The dynamic spread function is the critical element of the proposed strategy.
4. Derive the governing equation for *δ(t)*. Provide the required coefficient functions using published *T(t)* profiles for a baseline infection scenario. Systematically alter the baseline spread function, and solve the governing equations, to simulate alternative strategies for countering the infection.
5. Designate the time origin for the dynamic analysis as the point of the first intervention into the epidemic. For CNYC, we identify this as day 17 (from the first reported infection), when shelter-in-place was instituted. Prior to that point, it is assumed that *δ* is constant in time, and a traditional SIR model applies. The parameters for the traditional SIR model can be estimated from the published growth rate and reproduction number. The resulting values serve as initial conditions for the dynamic-spread-function analysis.

### 2.2 Development of Governing Equations

The model is based upon a 4-equation set consisting of 3 standard SIR equations for the susceptible, infected, and removed populations, plus an additional relation to describe droplet dynamics. The set was introduced by Stilianakis and Drossinos (2010) and extended by Myers et al. (2016) to explicitly account for the influence of protective equipment. We introduce the equations using a notation in which the primes denote dimensional quantities, with units such as numbers of persons or 1/day. The primes will be dropped following nondimensionalization. The basic set is as follows

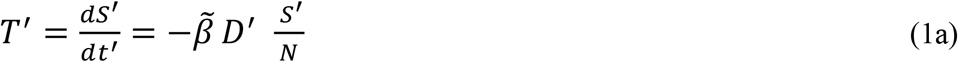

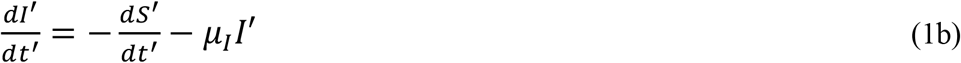

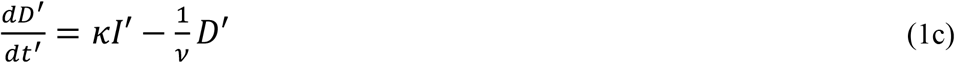

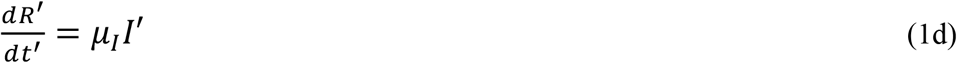

Here *S*′ is the number of susceptible individuals in the total population *N, I*′ is the number of infected individuals, *R*′ is the number of removed (recovered or died) individuals, *D*′ is the total number of droplets contributing to the spread of the disease, 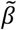 is the transmission rate, *μ*_*I*_ is the infection recovery rate, *κ* is the droplet production rate, and *v*^−1^is the droplet removal rate. We allow the transmission rate 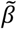 and the production rate *κ* to vary with time. The removed population is not of interest in the model, hence, the equation for *R*′ will not be considered further.

In the model applications, it is convenient to work with a nondimensional set of equations. Nondimensionalization of the dependent and independent variables, followed by arrangement of the resulting parameters in each equation in clusters, effectively reduces the total number of parameters. The resulting parameter combinations represent ratios than can lend insight into the infection dynamics. For example, the significance of a droplet production rate *κ* given in number of droplets per day can be difficult to appreciate, but the ratio *κ* /*v*^−1^of the droplet production rate to the droplet removal rate can help explain a rapid increase in the number of infection. In the nondimensionalization procedure we attempt to use scalings that represent order-of-magnitude estimates for the relevant variable. In that way, the values of the dimensionless parameters represent realistic estimates for the ratio of two competing effects in the infection scenario. Retrospective analyses possess the advantage that some of the representative scales can be obtained from the known (baseline) infection curves. Properties of the subsequent retrospective simulations can often be anticipated based upon parameter values derived from the baseline computations. This is not generally possible with forecasting models. Details of the nondimensionalization process are as follows.

We introduce the maximum number of new infections recorded per day (roughly 5000/day for CNYC) α for characterizing the infection quantities. For the relevant time scale, which we label Δ, we choose the time interval between the first intervention (roughly day 17, measured from the first reported infection, for CNYC) and the day when the number of new infections per day reaches a maximum (day 37 for CNYC). This time scale was chosen because it is representative of the interval over which the number of new infections changes by a significant (e.g. half the maximum) amount. Δ is 20 days for CNYC. The parameters *α* and Δ together characterize the infection scenario and can be used to nondimensionalize the system. When the population is large (e.g. New York City or New York State), it is convenient to work in terms of the difference between the susceptible population *S*′ and the total population *N*, because throughout the epidemic the entire susceptible population deviates only slightly from *N*. We call this difference population 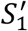 and set 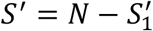. The relations used to relate the dimensional (primed) variables to the nondimensionalize (unprimed) variables are:

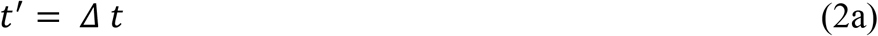

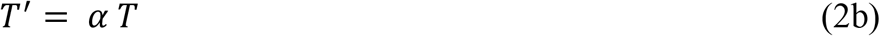

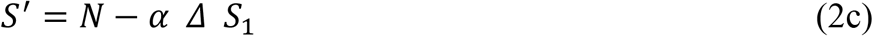

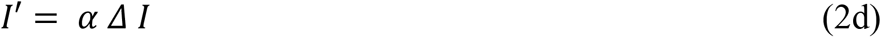

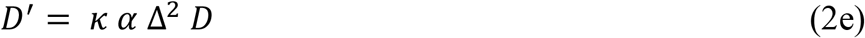

Using these relations in Equations (1a – 1c) and combining terms yields:

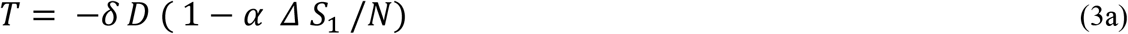

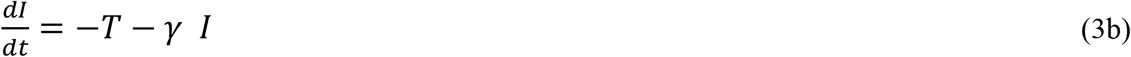

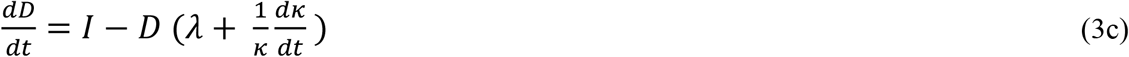

where *γ*=*Δμ*_*I*_ is the dimensionless infection recovery rate,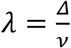 is the dimensionless droplet removal rate, and

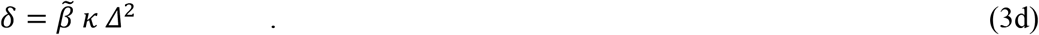

Since 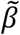 and *κ* vary with time, *δ* is also a function of time. As noted above, we designate *δ(t)* the “spread function”.

We apply the model only to large-population scenarios, where (*α Δ*) ≪ *N*. For CNYC, this inequality is well satisfied throughout the course of the COVID-19 epidemic. In that case, the last term in Eq. (3a) (*α Δ S*_1_/*N*) can be ignored. Additionally, the last term in Eq. (3c), which can be written as 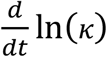, is considerably smaller than *λ*. In dimensional terms (removing the time scale Δ from *dt* and *λ*), 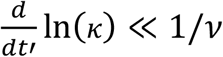, i.e. the rate of change of the logarithm of the droplet production is much smaller than the rate of droplet removal from all sources (droplet inactivation, inhalation, filtration…) Results from the computations featured in Section 3 showed that the rate of change of the logarithm of the production rate is on the order of 0.01/day. A range of removal rates was considered in our calculations; all were on the of 1/day. Ignoring the final terms in equations (3a) and (3c) gives

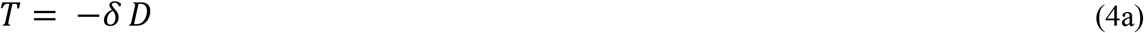

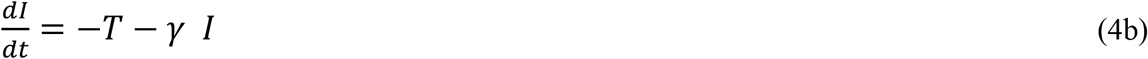

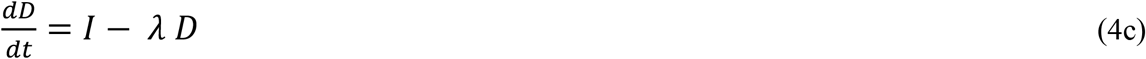

Using Eq. (4a) for *T* in Eq. (4c), and carrying out the differentiation and multiplying by *δ*, gives the two-equation system:

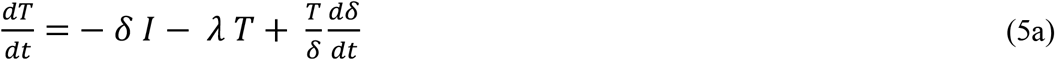

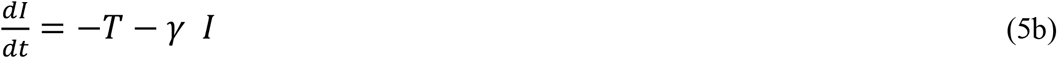

As noted above, we take the origin to be the time of first intervention.

To determine the dynamic spread function for the baseline scenario, we reformulate Eq. (5a) as an equation for *δ(t)*, assuming *T(t)* and *I(t)* to be known. The *T(t)* profile is obtained from the published number of new infections per day in the locale of interest (e.g. New York City). We label this published profile *T*_*b*_*(t)*, where the “b” subscript denotes baseline, and the resulting (from Eq. (5b)) infection profile *I*_*b*_*(t)*, and we insert them into the equation for *δ(t)*. The resulting equation for the baseline dynamic spread function is:

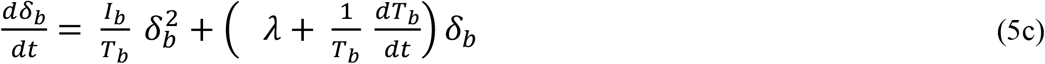

Because the governing equation for *δ*_*b*_*(t)* is informed by the published *T*_*b*_*(t)* profile, solving Eq’s (5a,b) using this dynamic spread function will reproduce (within numerical tolerances) the published *T*_*b*_*(t)* curve. The utility of *δ(t)* derives from modifying it to model alternative intervention strategies and solving Eq’s (5) to determine the new infection curves (i.e., *T(t)* and *I(t)* profiles). Modifications to account for protective strategies were performed in the following manner.

### 2.3 Accounting for Protective Equipment

We build upon a previously developed SIR model (Myers et al. 2016, Yan et al. 2018) that systematically accounts for the presence of protective equipment. Assuming both 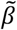 and *κ* vary with time, we can write (Eq. (3d))

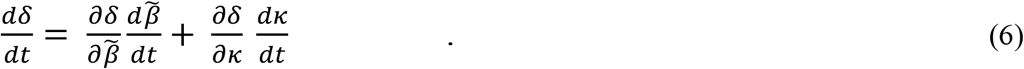

We apportion a fraction ϵ_κ_ (e.g. 1/5) of the change in *δ* to changes in droplet production, and accordingly set

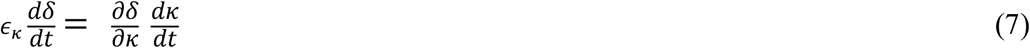

Since from Eq. (3d)

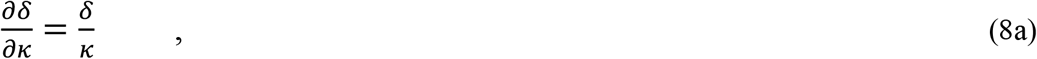

then

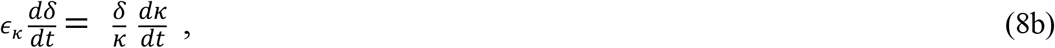

which can be integrated to

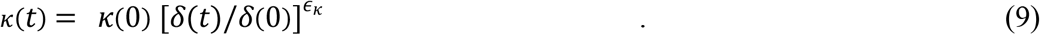

In Myers et al. (2016), it was shown that the droplet production rate in the presence of protective equipment can be written as

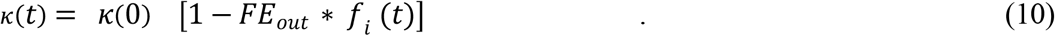

Here *FE*_*out*_ is the filtration efficiency (e.g. the *FE* for an N95 respirator is 95%) of the mask against outward-going particles produced by the infected individual wearing the mask (also referred to as source control). If *FE*_*out*_ varies with particle size, we assume that a dominant particle size exists in the distribution generated by the infected population, and the *FE* for that size applies. The quantity *f*_*i*_ is the fraction of the infected population wearing the mask at any given time, also known as compliance rate. We assume that the infected population wearing masks includes both symptomatic and asymptomatic persons, and that the masks have an equal effect on reducing the droplet production rate of symptomatic and asymptomatic persons. Eqs. (9) and (10) can be combined to give

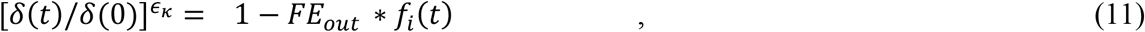

and

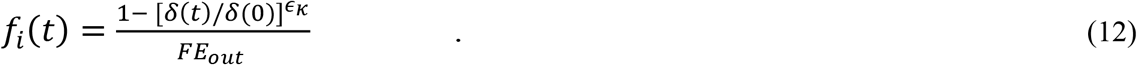

For a given scenario with spread function *δ*_*b*_*(t)* (derived from Eq. (5c)) and a given baseline mask material (i.e. *FE*_*out,b*_), Eq. (12) allows the population compliance rate *f*_*i*_ to be determined as a function of time, once the relative importance of change in production (ϵ_κ_) compared to change in transmission (1-ϵ_κ_) is estimated. Thus, to perform a retrospective analysis in which a barrier material of different outgoing-particle-capturing efficiency is investigated, the baseline compliance rate as a function of time would first be determined from Eq. (12), then that compliance profile and the new *FE*_*out*_ value would be used in Eq. (10) which, with Eq. (9), would be used to create a new dynamic-spread function. The modified spread function would then be used (in Eqs (5a,b)) to estimate the change in infection rate. A detailed example is provided in Section 3.1.

### 2.4 Solution Technique

The initial conditions for Equations (5) are obtained by first simulating the dynamics of the infection prior to any intervention, e.g. days 1 – 17 for CNYC. In that case, the derivative of the dynamic spread function is zero and Equations (5a,b) revert to a traditional SIR model. Seeking solutions that have an exponential time dependence of the form *exp*(Mt) for *T(t)* results in the algebraic equation

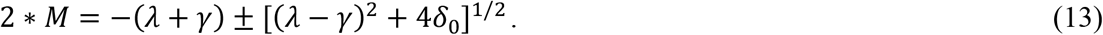

The subscript “0” on δ implies that the value applies to the initial period of the infection, before intervention occurs. The growth rate *M* can be obtained from infection rates published during the beginning of the epidemic, prior to any intervention, e.g. days 1 - 17 in CNYC. The other parameters do not vary during the course of the epidemic and are not subscripted. An exponentially growing solution will occur when

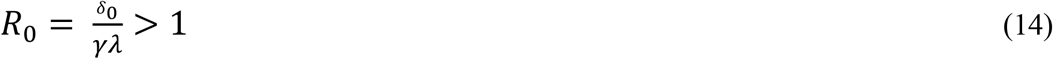

The symbol *R*_*0*_ represents the reproduction number (Myers et al. 2016) for the standard SIR model. Estimates of *R*_*0*_ for the early stages of epidemics are also published. In the simulations we perform, a range of recovery times *μ*_*I*_ (with corresponding dimensionless recovery times γ) ranging from 2 days to 10 days was considered. For any given value of γ, λ and δ_0_ were obtained using published values of the reproduction number *R*_*0*_ (Eq. (14) and growth rate *M* (Eq. (13)) for the scenario of interest. The *T*_*b*_*(t)* profile for CNYC was obtained from the Johns Hopkins Coronavirus Resource Center, wherein the data beginning at day 17 was used. *I*_*b*_*(t)* was derived from *T*_*b*_*(t)* using Eq. (3b), rather than using a published infection profile, so that it was not necessary to ascertain how well recoveries were tabulated in the published infection curves.

Uncertainties were determined by performing simulations for an ensemble of (*μ*_*I*_, *R*_*0*_) combinations, with each parameter selected from the range of published values for a given scenario. Six parameter sets were typically used to determine the uncertainty. The mean (over the six-parameter ensemble) *T(t)* profile, as well as a standard deviation above and below the mean at each instance of time, are reported.

In the simulations performed to examine different alternate intervention strategies, baseline *δ(t)* profiles using the actual intervention strategy were first obtained, and then modified to reflect alternatives. The governing equations (5 a,b) containing the modified spread function were solved using a Runge-Kutta method (Matlab *ode45*, Mathworks Inc.).

## 3. Results

We performed a variety of retrospective analyses involving protective equipment. For CNYC, days 17 - 37 were analyzed. This interval was chosen because day 17 is the day of the first intervention (shelter in place), and day 37 is the time of maximum new infections per day, based upon a 7-day average (Johns Hopkins Coronavirus Resource Center 2020). Initial reproduction numbers between 2 and 6 were considered, along with recovery times between 2 days and 10 days. The fraction ϵ_κ_ required to determine the compliance (Eq. (11)) was assumed to be 1/5. The infection scenario involving the entire state of New York was also considered in another set of simulations.

### 3.1 Effect of Mask Efficiency in CNYC

Using the procedure described in Section 2.3, we analyzed scenarios where the infected population in New York City deployed different types of masks. It was assumed that only the infected population deployed the masks, i.e. we considered a source-control measure. To illustrate the procedure of Section 2.2 for this scenario, we identify the outward filtration efficiency of the baseline mask as *FE*_*out,b*_, where the “b” subscript denotes baseline, and the outward filtration efficiency of the modified mask design as *FE*_*out,mod*_. Using the 7-day average data (Johns Hopkins Coronavirus Resource Center 2020) for CNYC in Eq. (5c) generates the baseline source function *δ*_*b*_*(t)*. The baseline mask compliance profile *f*_*i,b*_ (Eq. (12)) is

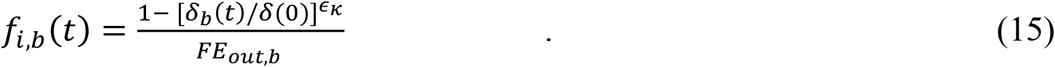

We assume the compliance profile for the modified mask strategy is identical to baseline, i.e. the population uses a higher-efficiency mask but with the same adoption rate. Using the modified mask filtration rate *FE*_*out,mod*_, along with the baseline compliance profile (Eq (15)), in Eq. (10) for the droplet production gives the modified droplet production rate

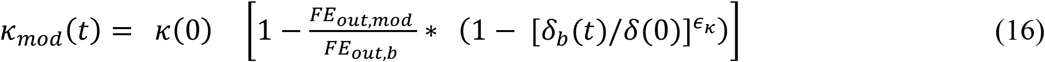

Since the transmission rate 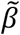 does not change in this retrospective simulation, from Eq. (3d) we can conclude

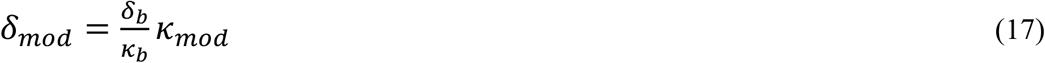

Using Eq. (9) to prescribe the baseline production rate and Eq. (16) to prescribe its modification gives

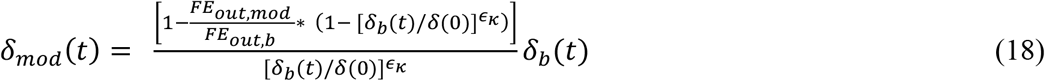

This expression used as the spread function in Equations (5 a,b) enables simulation of scenarios involving masks of different filtration efficiencies. For baseline, the filtration efficiency was taken to be 67%. This value is representative of homemade masks (Howard et al. 2020), though the filtration capability of homemade masks spans a wide range. For the modified scenarios, higher-efficiency masks with FE’s of 75%, 80%, and 90% were considered.

Figure 1a shows the dynamic spread function as a function of time for CNYC. A sharp decrease is seen initially, owing to the shelter-in-place restriction. For larger FE, a slightly sharper decrease in the dynamic spread function is observed. A slight decrease in dynamic spread function value is associated with a much larger decrease in new infections (Fig. 1b). Increasing FE from 67% to 75%, for example, reduces the dynamic spread function value a few percent at day 37, while the maximum number of new infections (at day 37) decreases by about 15%. The turn-around time is decreased from about 37 days to 35 days.

**Figure 1a.**
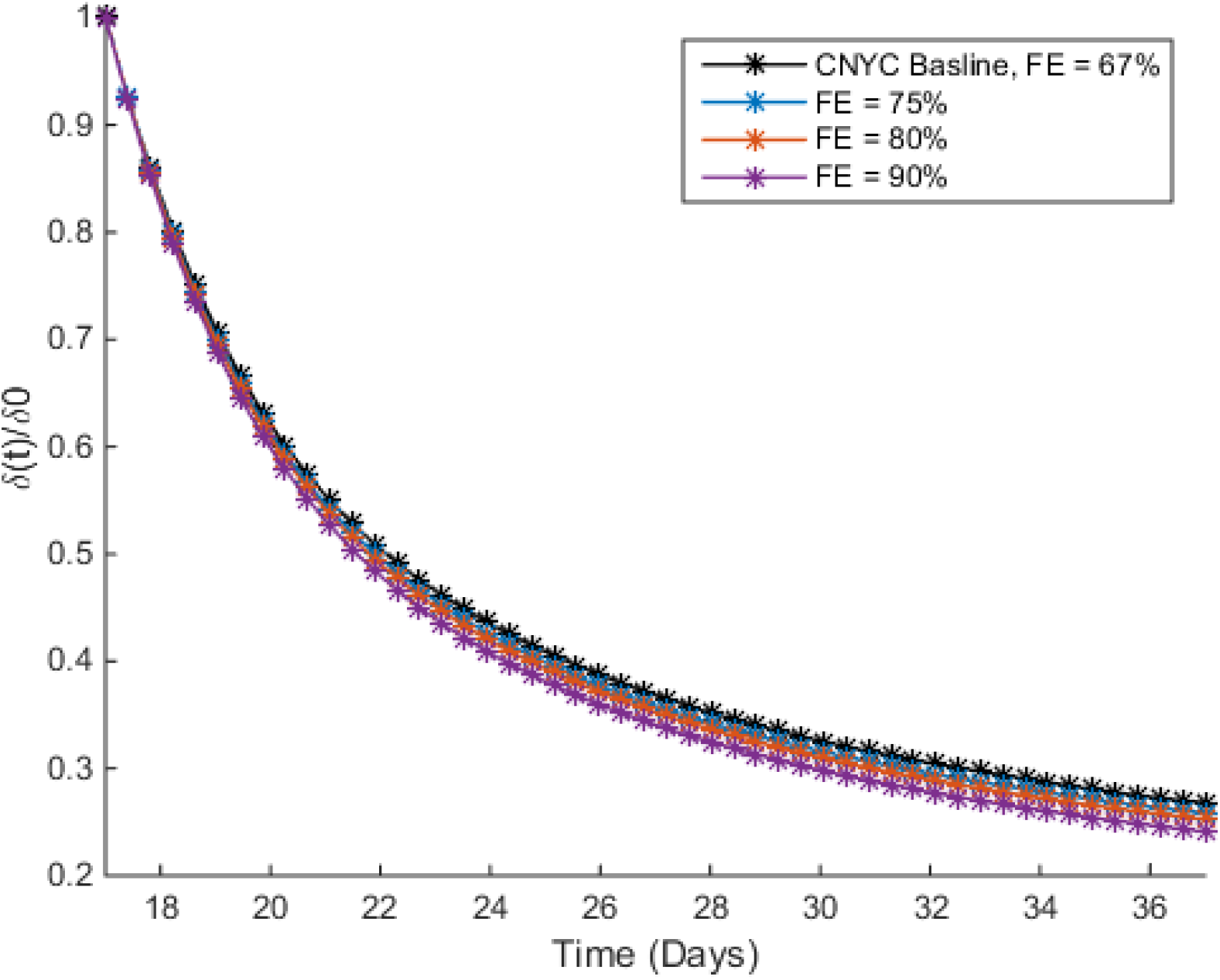
Dynamic spread function for CNYC with infected population deploying masks with different filtration efficiencies (FE).

**Figure 1b.**
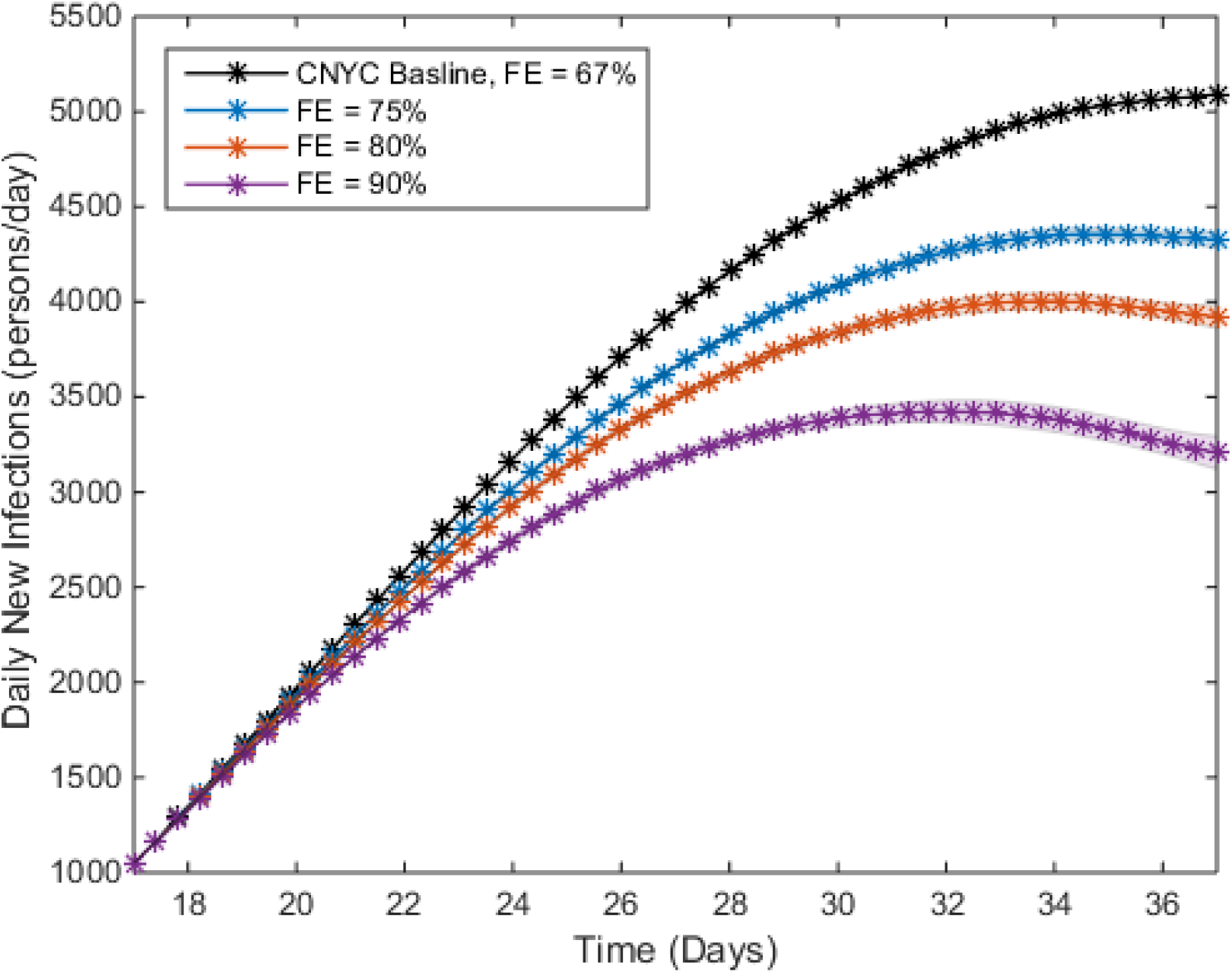
Number of new infections per day for CNYC, with infected population deploying masks with different filtration efficiencies (FE). Shadowed regions denote values within a standard deviation of the mean, for an ensemble of simulations using different reproduction numbers and recovery rates.

For the same increase of FE from 67% to 75%, the number of infected individuals (Fig. 1c) at day 37 is reduced by about 30%. The uncertainty is considerably larger for the infected population (Fig. 1c) than the number of new infections per day (Figure 1b), because the infected population is much more strongly influenced by the recovery time than the number of new infections. The recovery time spanned a factor of 5 over all the simulations performed. The uncertainty for the dynamic spread function (Fig 1a) is comparable to that for the infected population, though for clarity it is not shown.

**Figure 1c.**
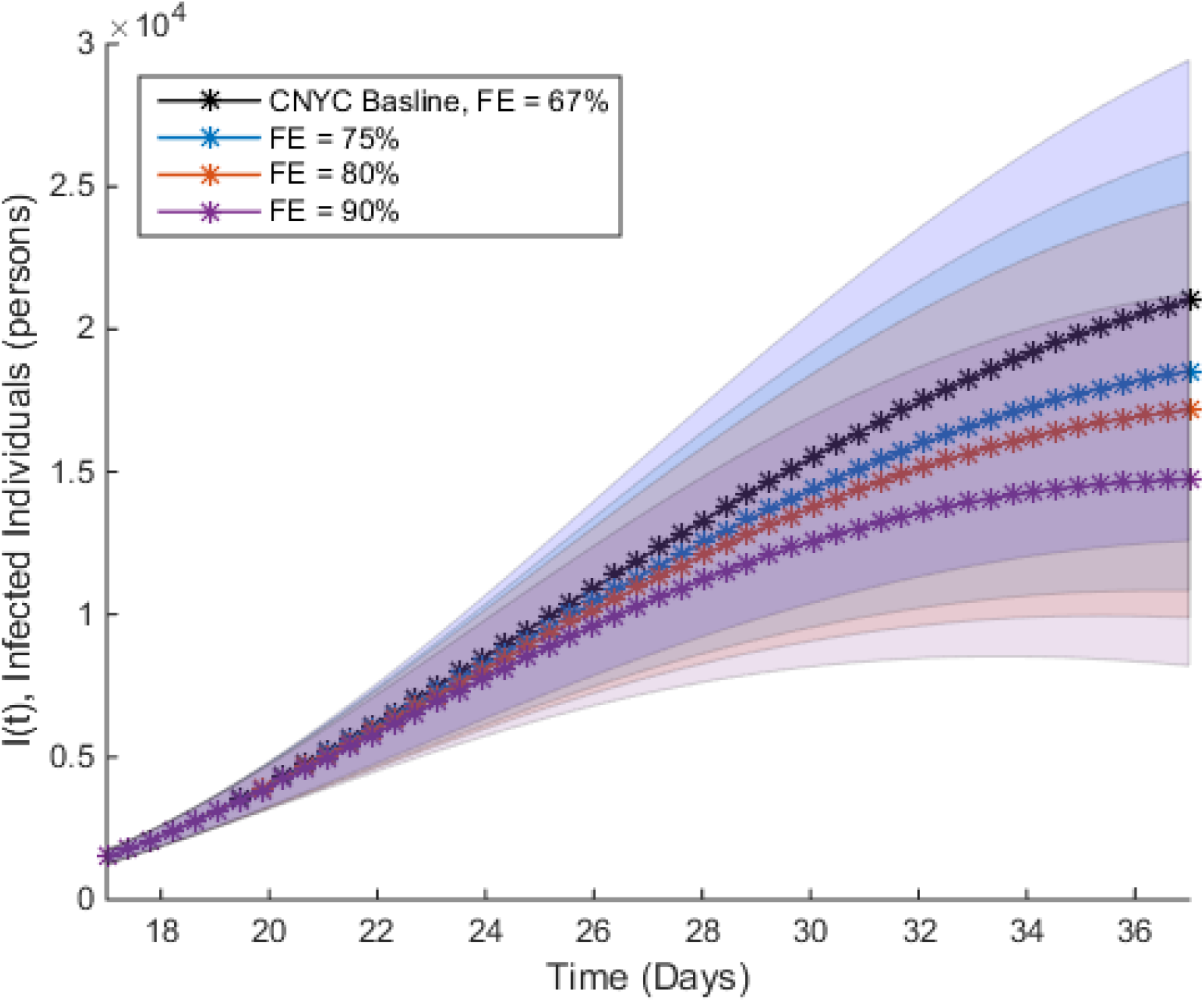
Infected population as a function of time for CNYC, when infected population deploys masks of different filtration efficiencies (FE). Shadowed regions denote values within a standard deviation of the mean, for an ensemble of simulations using different reproduction numbers and recovery rates.

### 3.2 Effect of Mask Compliance in CNYC

To evaluate the effect of mask compliance, we assume that the adoption rate for the mask follows a temporal profile identical to baseline, but larger or smaller by a factor of *F*. As in the previous set of calculations, it is assumed that only the infected population deploys the masks. Baseline compliance as a function of time is again given by Eq. (15), and the compliance profile for the modified scenarios is *F* times this expression. Using this modified compliance in Eq (10) gives the modified production rate

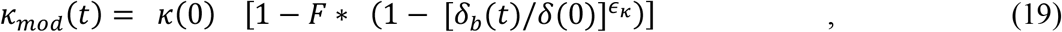

analogous to Eq. (16) for the variable filtration-efficiency simulations. The baseline spread function *δ*_*b*_(*t*) is identical to that for the filtration-efficiency study; it is given by the top curve in Fig. 1. As in the variable-FE case, the transmission term is not altered in the variable-compliance simulations. Thus, the steps used to generate the spread function (Eq. (18)) from the production term (Eq. (16)) can be repeated here, with Eq. (19) replacing Eq. (16) as the representation for *κ(t)*. The result is

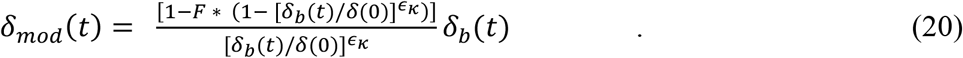

For the mask compliance analysis, a mask of FE of 67% was assumed. Upon solving for *f*_*i*_*(t)*, (Eq. (15)) it was found that the baseline fraction *f*_*i*_*(t)* of the CNYC population wearing the mask increased from 0% at day 17 to about 42% on day 37. The 42% maximum compliance was increased to 50%, 60%, and 70% in a series of computations by adjusting the *F* value in Eq (20), and solving Eqs (5a,b) with this modified spread function. *F* was iteratively adjusted to yield the target compliance at day 37.

Increasing the mask compliance to 50% reduced the maximum number of new infections per day from about 5100 to 4000 (Fig. 2). The turn-around time is reduced from approximately 37 days to 34 days. Similarly, increasing the mask compliance from baseline to 60% and 70% reduced the number of new infections per day by 42% and 58%, respectively. The corresponding turn-around times reduced to 31 days and 28 days (Fig. 2).

**Figure 2.**
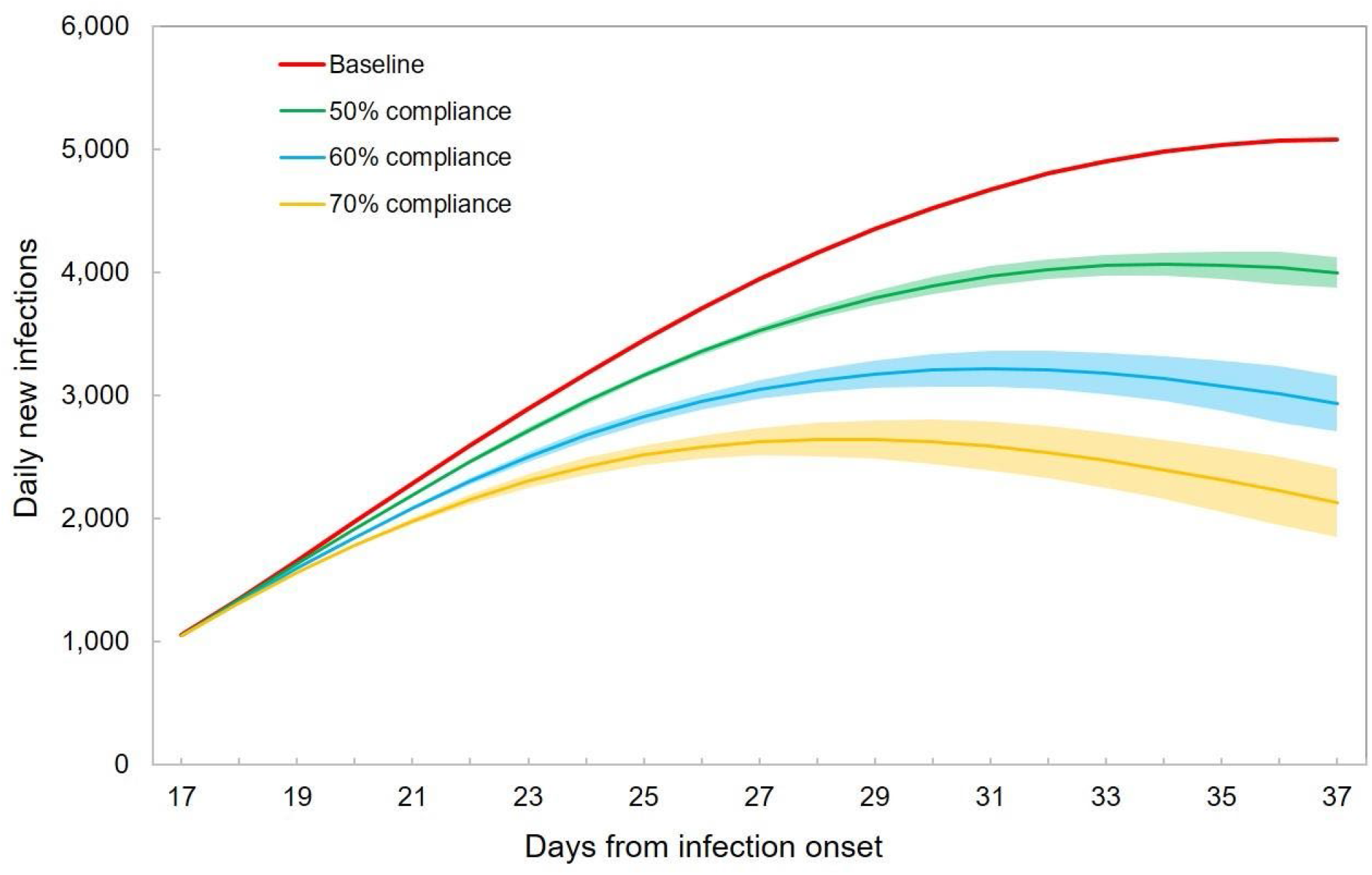
Number of new infections per day for scenarios where various fractions of the infected population in CNYC deploy masks with a 67% filtration efficiency.

### 3.3 New York State Scenario

To compare the model’s determination of population behaviors in New York City versus that of the New York State, and to compare with results computed by other investigators, we briefly consider the COVID-19 scenario in New York State. Infection data for the state was obtained from New York Times Github Database (2020). For New York State, days 23 – 38 were analyzed. This interval was chosen because day 23 is the day when the first lockdown order was executed (ABC News Report, 2020), and day 38 is the time of maximum new infections per day in the first infection wave, based upon a 7-day average (New York Times 2021).

The baseline dynamic spread function *δ*_*b*_*(t)* for New York State was determined from Eq. (5c), in an equivalent manner to the CNYC case. In this brief comparison of New York City with New York State, only one combination of *μ*_*I*_ (or γ) and *R*_*0*_ values (representing the lower end of the range of published values) was considered.

The baseline dynamic spread functions for New York City and New York State are plotted in Figure 3a. The initial decay in the spread function is steeper for New York State. This translates into considerably fewer new infections (relative to the total population), and a shorter turn-around time, as seen in Figure 3b.

**Figure 3a.**
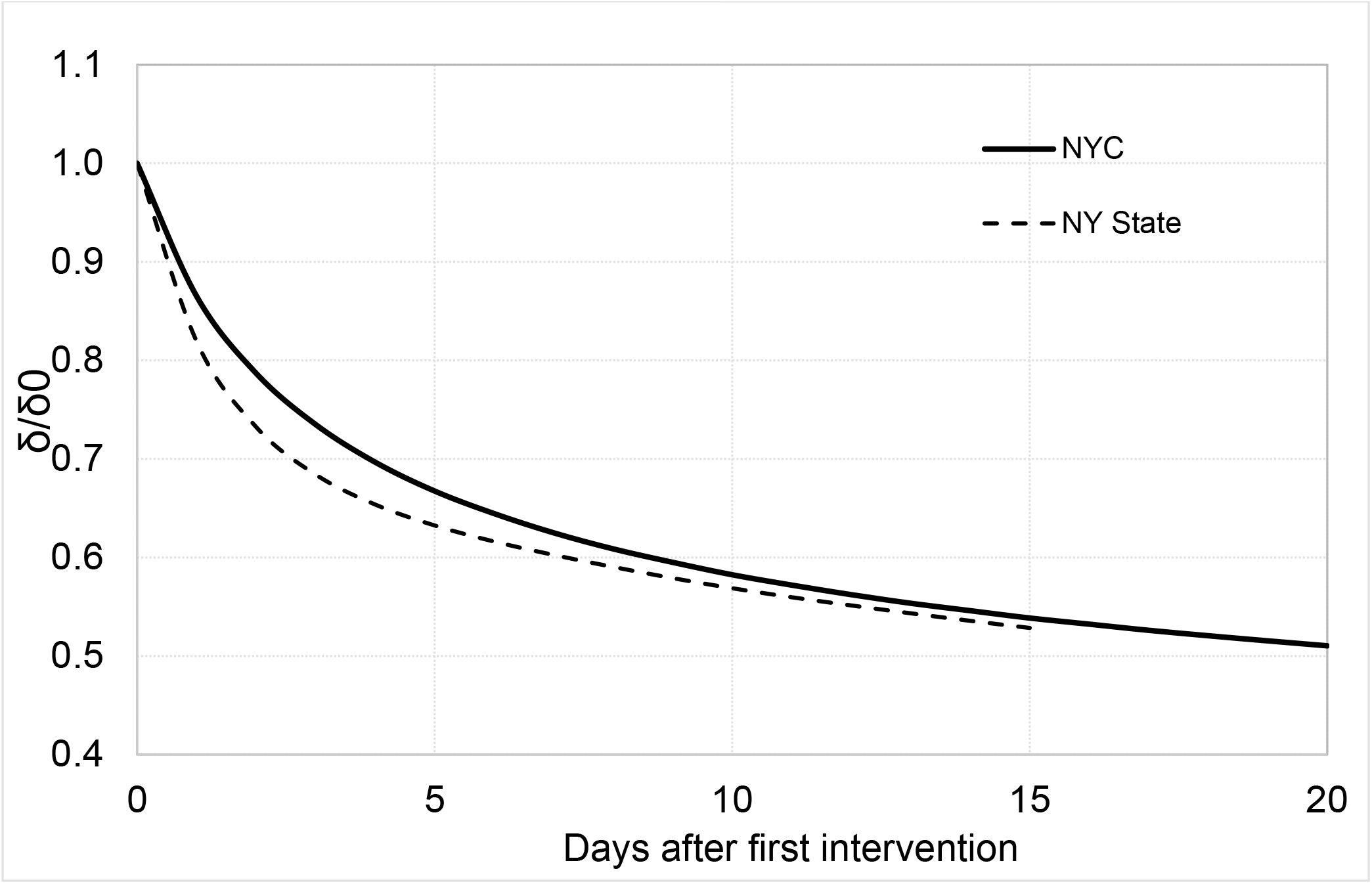
COVID-19 dynamic spread function as a function of time after first intervention, for both New York City (solid) and New York State (dashed).

**Figure 3b.**
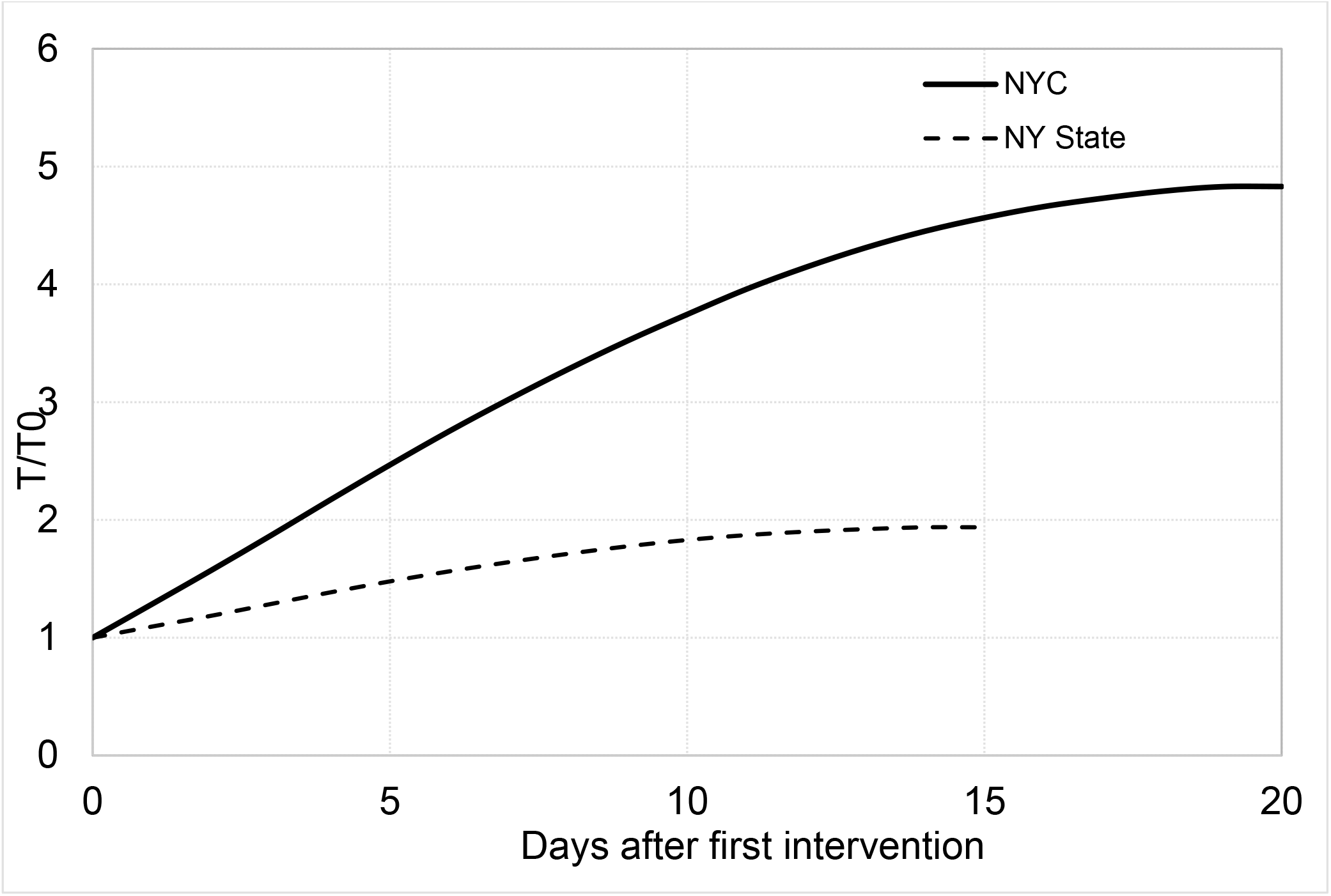
Numbers of new COVID-19 infections in New York City (solid) and New York State (dashed), as a function of time after the first intervention.

We note that the values plotted in Fig. 3b are just baseline infection curves. No alternative interventions were involved.

The New York State simulation is useful for comparison with the study by Ngonghala et al. (2020), who also analyzed alternative COVID-19 intervention strategies involving protective equipment and social-distancing measures. Ngonghala et al. (2020) computed numbers of deaths and hospitalizations in New York State as the level of social distancing and type of mask and mask compliance were varied. For the comparison, we assume that our primary dependent variable, numbers of new infections, is proportional to the hospitalization rate throughout the time range of interest (days 23 – 38 in New York State).

To incorporate variable social distancing into the dynamic spread model, modifications to the spread function were made in the following manner. The level of social distancing is related to the transmission rate 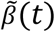. As described in Stilianakis and Drossinos (2010) and Myers et al. (2016), the transmission rate is proportional to the number of contacts between a susceptible individual and an infected person. Thus, the level of social distancing is proportional to 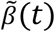. This function is similar to the effective contact rate in Ngonghala et al. (2020). Since the dynamic spread function *δ(t)* is proportional to the transmission rate 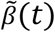 (Eq. (3d)), different levels of social distancing could be implemented in a straightforward manner by scaling the baseline spread function (dashed curve in Fig. 3a) by the desired amount. That is,

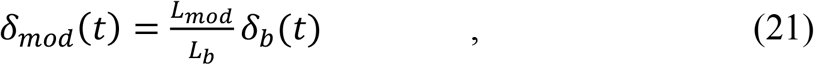

where *L*_*b*_ is the baseline level of social distancing and *L*_*mod*_ is the modified level of social distancing, and *δ*_*b*_(*t*) is the baseline spread function for New York State. Solving Eqs (5 a,b) with the scaled spread function yielded the infection curves for the modified level of social distancing. Effects of mask efficacy and compliance rate in the New York State scenario were determined by the same procedure used to generate the CNYC results in Figs 1 and 2.

The dynamics for COVID-19 spread in New York State as a function of social distancing level, as predicted by Ngonghala et al. (2020) and the dynamic-spread model, are presented in Table 1. For each model, the percent reduction (relative to baseline) in the maximum number of new infections or hospitalizations is tabulated for different reductions in social distancing (i.e. reductions in β). For the dynamic-spread model, when the level of social distancing was reduced by more than 30%, no maximum was attained after the initial day (exponentially decaying number of new infections following intervention), so the comparison was made at day 38, where the baseline maximum was attained.

**Table 1.** COVID-19 Dynamics for New York State for Different Levels of Social Distancing

| Reduction in Level<br>of Social<br>Distancing (%) | Reduction in Maximum<br>Number of New Infections<br>(%) | Reduction in Maximum Number of<br>Hospitalizations Predicted by Ngonghala<br>et al., 2020 (%) |
| --- | --- | --- |
| 10 | 35 | 24 |
| 20 | 48 | 48 |
| 30 | 85* | 72 |
| 40 | 92* | 92 |

\*No maximum at this level of social distancing; comparison performed at day 38

A larger reduction in the infection metric (number of new infections or number of hospitalizations) is predicted by the dynamic-spread model for the lowest reduction in level of social distancing, but otherwise comparable reductions are predicted by the two models.

COVID-19 infection metrics in New York State as a function of mask efficiency and compliance are presented in Table 2. As with social distancing, high levels of mask efficiency and compliance yield exponentially decaying rates of new infections after the first intervention. Hence, comparisons are made at day 38, the time of maximum number of new infections for the baseline case. When the product of the mask efficiency times compliance is low, the dynamic - spread model predicts a larger decrease (up to a factor of 1.7) in the infection metric (new infections or hospitalizations) than the model of Ngonghala et al. (2020). Otherwise, comparable changes in the infection metric are predicted by the two models when different mask strategies are implemented.

**Table 2.** COVID-19 Dynamics for New York State for Different Mask Efficiencies and Compliance

| Final Compliance $f_i$ (%)<br>($c_m$ in Ngonghala et al., 2020) | Reduction in Maximum<br>Number of New<br>Infections (%) | Reduction in Maximum Number of<br>Hospitalizations Predicted by<br>Ngonghala et al., 2020 (%) |
| --- | --- | --- |
| FE = 25% |  |  |
| 10 | 15 | 8.7 |
| 25 | 34 | 22 |
| 50 | 56 | 43 |
| 75 | 72 | 64 |
| FE = 50% |  |  |
| 10 | 28 | 19 |
| 25 | 56 | 44 |
| 50 | 75 | 82 |
| 75 | 93* | ~100 |
| FE = 75% |  |  |
| 10 | 39 | 26 |
| 25 | 69 | 63 |
| 50 | 93* | ~100 |
| 75 | 98* | ~100 |

\*Comparison Performed at Day 38

## 4. Discussion

To the extent that reduction in the spread of infection is due primarily to reductions in social distancing compared to lower rates of droplet production, i.e. assuming ϵ_κ_ < < 1, then the spread function curve (top line, Fig. 1a) represents the level of social distancing in New York City during the COVID-19 crisis. Within the first 5 days after the stay-at-home order, the level of social distancing drops by half. A slower adoption of the stay-at-home order is observed after that, but by the turn-around point (day 37), another factor-of-two reduction in social-distancing is achieved. These trends illustrate the continuous nature by which interventions take place during epidemics. The state of New York adopted the required behavioral changes faster than the city of New York (Fig. 3a). This faster rate of behavior modification resulted in a considerably smaller number of new infections (relative to the total population) in New York State. Besides enabling the simulation of alternative intervention strategies, the dynamic function in Fig 3a is useful in interpreting the baseline trends in Fig. 3b.

Another useful interpretation of the dynamic spread function can be acquired by writing the function (Eq. (3d)) as 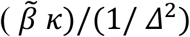. The top, with dimensions of (1/time)^2^, i.e. (rate)^2^, can be interpreted as the rate at which quantities promoting the spread of the infection are produced and transmitted. The denominator, also with units of (1/time)^2^, is inversely proportional to (the square of) the time over which the population responds to the infection. *δ*^1/2^, then, can be thought of as the ratio of the infection spread rate to the societal response rate. The relative intensity of epidemics at different locations, or in a given population at different times, can be characterized by the different values of the square root of the dynamic spread function.

The changes in infection metric (number of new cases or number of hospitalizations during the COVID-19 epidemic in New York State) due to a variety of interventions involving social distancing and mask usage were computed using the dynamic-spread model and the model implemented by Ngonghala et al. (2020). The two models agreed within about 25% on average. This was felt to be a relatively small difference, given the large number of assumptions in SIR models. Some caveats are in order, though. First, it is unclear to what extent relative (to baseline) changes in new infections can be compared with relative changes in hospitalizations. Second, the time course for the hypothetical scenarios was quite different between the dynamic-spread model and that of Ngonghala et al. (2020). For example, in Ngonghala et al (2020), application of masks with higher FE than baseline resulted in curves that are flatter than baseline, while in the dynamic -spread model the curves largely retained the same shape (Fig. 2). This is due, at least in part, to the fact that behavioral changes occurred gradually in the dynamic-spread model, and they did not commence until the time of first intervention. In Ngonghala et al. (2020), and many other constant-parameter models, parameter values characterizing hypothesized scenarios with higher degrees of protection are implemented at the onset of the epidemic. This results in a flat infection profile. An additional difference between Ngonghala et al. (2020) and the dynamic-spread model is that we assumed only the infected population deployed masks. Adding mask usage by the susceptible population would increase the reduction in new infections. This would enlarge the difference between the dynamic spread model and that of Ngonghala et al. (2020) for most scenarios, since the dynamic-spread model usually showed a larger reduction. However, since the fraction of the reduction in the dynamic spread function attributed to masks was small (ϵ_κ_ = 1/5), incorporating mask usage by the susceptible population would not significantly affect the values in Tables 1 and 2. Mask usage by the susceptible population was not included in the dynamic-spread model, owing to the fact that the *FE* is often different for inward (susceptible people) and outward (infected people) flux of pathogens, and further analysis was thought to be necessary before assigning mask efficiencies appropriate for the susceptible population.

While days 17 to 37 were featured in our simulations of CNYC, the dynamic -spread - function technique can be applied to any time interval where reliable numbers of new infections are available. The standard SIR model is used prior to the time when either the production rate or the transmission rate is altered by an intervention strategy. At that point the dynamic simulations commence, with the results from the standard SIR model serving as initial conditions.

The parameter ϵ_κ_, which quantifies the level of change in the dynamic spread function due to droplet production compared to that due to transmission, was chosen to be small (1/5) based upon the intuition that social distancing is more important than mask usage in altering infection dynamics. Other relatively small values of ϵ_κ_, e.g. 1/3, yielded similar results to those reported above. The ϵ_κ_ parameter is useful for estimating the mask compliance (Eq. (12)), as well as the level of social distancing, as a function of time. Alternatively, if information is available on the compliance profile for the scenario of interest, it can be used in Eq. (12) to determine ϵ_κ_ more rigorously.

The effects of different protective-equipment strategies in New York City and New York State were investigated without having to update the SIR-model parameters due to interventions during the epidemics. The continuous adoption of masks is difficult to simulate by updating coefficients at various times in standard SIR models. With the dynamic-spread approach, the gradual adoption of masks is captured in a natural manner.

For the conditions of the simulations, a slight increase in facemask efficiency resulted in a larger benefit than a commensurate increase in compliance. At day 37, for example, a fractional increase in compliance of 0.1 resulted in a reduction in new infections of about 500 per day (Fig. 2), while a fractional increase in *FE* of 0.1 reduced the number of new infections by about 800 (Fig. 1b). For a higher baseline *FE*, increasing the compliance would produce a larger decrease in new infections. This comparison between filtration efficiency and population compliance illustrates the utility of the model for determining how resources devoted to countermeasures can be optimally spent. In this case, the model can help inform the choice between 1) producing and distributing barriers of higher FE, and 2) educating and incentivizing the population to deploy barriers more readily available.

The model is not intended to be a prediction tool, in the sense of forecasting the future course of an ongoing epidemic. The purpose of the model is to compare different intervention strategies for scenarios where the baseline infection profile (number of new infections per day) is provided. Also required are the initial reproduction number and an estimate of the recovery rate. Though the model is not a forecasting tool, it can be useful for designing future countermeasures (e.g. for successive waves of an epidemic), particularly if elements of the anticipated scenario are similar to those of the baseline scenario used to compute the dynamic spread function *δ(t)*. These elements include, most importantly, population behaviors such as face mask adoption (affecting both *κ* and 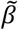 in Eq. 5) and social distancing (affecting 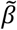), but also environmental factors such as the pathogen inactivation rate. We refer the reader to Stilianakis and Drossinos (2010) for the dependence of infection dynamics on the numerous properties of the pathogen, the population, and the environment. Here we emphasize that the dynamic spread function implicitly captures the influence of all these factors, even though no functional dependence of the parameters is introduced. Only when considering an alternative scenario that varies one of the factors (e.g. the production rate *κ* in this study) does the explicit parametric dependence enter. An important consequence of this property is that social distancing, likely the dominant factor affecting infection dynamics, was captured in the CNYC study without having to be explicitly modeled.

The method by which the dynamic-source model simulates changes in population behavior differs from the manner in which it is typically done with SIR models in two important ways. With most SIR models, changes in population behavior are addressed by updating parameters at discrete times. However, the original set of parameters and parameter updates is not unique. Depending upon the strategy used by an investigator to minimize differences with published infection rates, and the “best guesses” made for the parameters that aren’t well informed by data, the investigator can derive significantly different parameter values from another investigator using the same equations, even when both investigators show good agreement with published infection curves. When the two investigators perform retrospective analyses in which a single parameter is varied, the results of the analyses can be sensitive to the baseline parameter values, which can vary for the two investigators. The dynamic-source method bypasses this potential uncertainty. The second important difference between the dynamic-source model and traditional SIR approaches regarding modeling behavior dynamics is the dynamic-source method treats the variation in population behavior over time as a mechanism affecting the infection dynamics. This mechanism is described by the last term in Eq. (5a). Even if a traditional SIR model updates the parameters periodically, this term is not included. The mechanism is analogous to transport in physical systems (e.g. fluids), where there is temporal or spatial variation in a property (e.g.viscosity). The gradient of the property multiplied by the transported entity (e.g. momentum) is a transport mechanism that should be included to fully capture the effects of property variation, in addition to simply updating the variable property at different times or spatial locations.

As noted above, the lack of forecasting ability is a limitation of the model. Similarly, because of the model’s simplicity, it cannot quantify the effects of the different factors affecting the infection dynamics, e.g. the roles played by symptomatic vs. asymptomatic infected persons. While, as mentioned above, any complicated mechanism that affects either the droplet generation (parameter *κ* in Eq. (5c)) or the infection transmission (parameter 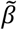 in Eq. (5c)) is automatically (i.e. as part of the calibration process, with no user input involved) included in the spread function during the calibration process, these effects are integrated with all other effects. If it is desired to explore how an intervention involving a particular mechanism would retrospectively alter the infection rate, a direct connection between that mechanism and either the droplet generation or the infection transmission would need to be specified. While that may not be possible given the model’s level of simplicity, we note that in future generations of the model an enhanced level of specificity should be possible. For example, in our calculations, all of the types of facemasks used in New York City were averaged together into a single representative barrier. If information is available to prescribe the levels at which different types of masks were deployed, it is likely that a modified spread function containing a sum over different mask designs weighted by their popularity could be constructed.

Like previous studies (Stutt et al. 2020, Eikenberry et al. 2020), our simulations predict that considerable benefit can be obtained from higher FE masks without requiring N95 levels of efficiency (Fig 1). It is important to emphasize that for the benefits to be realized, the FE for the barrier material must be attainable for the particle-size range of the dominant transmission mode for the given scenario. One way of assuring this is for the barrier material to provide the given FE across the spectrum of particle sizes. Otherwise, knowledge of the material FE for the intended application (e.g. reducing airborne particulates generated by coughing or sneezing by infected persons indoors) is required in order to generate useful estimates. The complex issues of dominant transmission mode for COVID-19, and the FE of different masks designs for the different modes, will be addressed in future applications of the model.

## Data Availability

No human, animal, or in-vitro data is included in the manuscript, which describes a risk-assessment model. Algorithms used to derive the mathematical results have been referenced.

## Author Contributions

JO and GD developed the computer code and performed the simulations. SB helped derive the governing equations and generated analytical solutions used to verify computations. SBB researched COVID-19 databases and performed operations to optimize data use by the model. MM designed the study. All authors contributed to the writing and editing of the manuscript.

